# Altered kidney function and acute kidney damage markers predict survival outcomes of COVID-19 patients: A prospective pilot study

**DOI:** 10.1101/2021.01.10.20249079

**Authors:** Mustafa Zafer Temiz, Ibrahim Hacibey, Ramazan Omer Yazar, Mehmet Salih Sevdi, Suat Hayri Kucuk, Gizem Alkurt, Levent Doganay, Gizem Dinler Doganay, Muhammet Murat Dincer, Emrah Yuruk, Kerem Erkalp, Ahmet Yaser Muslumanoglu

## Abstract

**Background:** The central role in the pathogenesis of severe acute respiratory syndrome coronavirus 2 (SARS-CoV-2), called as coronavirus disease 2019 (COVID-19), infection is attributed to angiotensin-converting enzyme 2 (ACE-2). ACE-2 expressing respiratory system involvement is the main clinical manifestation of the infection. However, literature about the association between the severe acute respiratory syndrome coronavirus 2 (SARS-CoV-2) infection and higher ACE-2 expressing kidney is very limited. In this study, we primarily aimed to investigate whether there is a kidney injury during the course of SARS-CoV-2 infection. The predictive value of kidney injury for survival was also determined.

**Methods:** A total of 47 participants who met the inclusion criteria were included in the study. The participants were classified as ‘‘COVID-19 patients before treatment’’ ‘‘COVID-19 patients after treatment’’, ‘‘COVID-19 patients under treatment in ICU’’ and ‘‘controls’’. The parameters comorbidity, serum creatinine and cystatin C levels, CKD-EPI eGFR levels, KIM-1 and NGAL levels, urine KIM-1/creatinine and NGAL/creatinine ratios were statistically compared between the groups. The associations between covariates including kidney disease indicators and death from COVID-19 were examined using Cox proportional hazard regression analysis.

**Results:** Serum creatinine and cystatin C levels, urine KIM-1/creatinine levels, and CKD-EPI, CKD-EPI cystatin C and CKD-EPI creatinine-cystatin C eGFR levels exhibited significant difference in the groups. The causes of the difference were more altered kidney function and increased acute kidney damage in COVID-19 patients before treatment and under treatment in ICU. Additionally, incidences of comorbidity and proteinuria in the urine analysis were higher in the COVID-19 patients under treatment in ICU group. Urine KIM-1/creatinine ratio and proteinuria were associated with COVID-19 specific death.

**Conclusions:** We found that COVID-19 patients under treatment in ICU exhibited extremely higher levels of serum cystatin C, and urine KIM-1/creatinine and urine NGAL/creatinine ratios. These results clearly described the acute kidney damage by COVID-19 using molecular kidney damage markers for the first time in the literature. Lowered CKD-EPI, CKD-EPI cystatin C and CKD-EPI creatinine-cystatin C eGFR levels were determined in them, as well. Urine KIM-1/creatinine ratio and proteinuria were associated with COVID-19 specific death. In this regard, considering kidney function and kidney damage markers must not be ignored in the COVID-19 patients, and serial monitoring of them should be considered.

## Introduction

Severe acute respiratory syndrome coronavirus 2 (SARS-CoV-2) infection, which is now called as coronavirus disease 2019 (COVID-19) by the World Health Organization, involves the respiratory tract as the primary target in most of the cases and has been known as the latest pandemic of the modern world [1, 2]. The main clinical manifestation of the disease ranges from asymptomatic course to severe pneumonia [2]. The central role in the pathogenesis is attributed to angiotensin-converting enzyme 2 (ACE-2). It enables viral entry into the target cells [3]. It is not surprising that SARS-CoV-2 easily settles down to one of the highest ACE-2 expressing tissues in the body and infects primarily respiratory system and lung which puts it into the respiratory system virus family [3, 4]. However, various tissues outside the lung have ACE-2 expression [3] including the kidney proximal tubule cells [3, 5].

In this context, the kidney is a potential target for SARS-CoV-2 infection. To date, there is no analytical study based on the assumption that the kidney is potential target of COVID-19. In the present study, we primarily aimed to investigate whether there is a kidney injury during the course of COVID-19 through a prospective analytical study with a pilot design. The secondary aim was identifying the predictive value of several kidney function parameters and damage markers for survival of COVID-19 patients.

## Materials and Methods

### Study design and setting, and participants

A prospective, pilot study was conducted at a tertiary center declared as pandemic hospital in Istanbul/Turkey by Turkish Ministry of Health (TMOH). A total of 75 patients with confirmed and probable COVID-19 diagnose and 11 healthy controls were prospectively evaluated between April and May 2020.

All the participants selected from urology and ICU departments of our hospital. Inclusion criteria were being more than 18 years old males and females, laboratory confirmed COVID-19 patients immediately after their treatment, highly suspected COVID-19 patients with specific computerized tomography (CT) imaging findings [6] before treatment and laboratory confirmed COVID-19 patients under the treatment in intensive care unit (ICU). Controls were selected from healthy individuals without any clinical or laboratory findings consistent with COVID-19. The presence of end-stage renal disease, previous kidney surgery history, presence of acute urinary tract infection, and presence of current urinary stone disease were the exclusion criteria. Patients with solitary kidney and urogenital malformation, and pregnant, puerperant and lactant patients were also not included in the study.

Diagnosis of the COVID-19 was confirmed on the nasopharyngeal and oropharyngeal swab samples with real-time reverse-transcription polymerase chain reaction (RT-PCR) in our genomic laboratory. We performed the RT-PCR according to manufacturer’s instructions (Coyote Bioscience Co., Ltd., Beijing, China). Suspected COVID-19 cases were determined using latest updated version of our national COVID-19 guidelines [7]. Suspected cases with specific lung CT findings [6] were termed as highly suspected COVID-19 cases. We again used latest updated version of our national COVID-19 guidelines to determine the treatment algorithms, as well. Initially, we stratified the participants into the four groups as ‘‘highly suspected COVID-19 cases before treatment’’, ‘‘confirmed COVID-19 patients after treatment’’, ‘‘confirmed COVID-19 patients under treatment in ICU’’ and ‘‘controls’’. Confirmed COVID-19 patients after treatment group was generated from the COVID-19 patients who were treated in the specific COVID-19 clinic and did not require ICU admission. We applied the recommended treatment algorithms immediately after the nasopharyngeal and oropharyngeal swab sampling, as the molecular test results required a few days which may result in delay of the treatment. Therefore, to create a further ‘‘confirmed COVID-19 patients before treatment group’’, highly suspected COVID-19 cases have been included even before their molecular test results had been provided. In this group, blood and urine samples were collected before starting to treatment. In the second group including confirmed COVID-19 cases with molecular test, sample collection was performed immediately after finishing the five days treatment course. In the third group, samples were collected at fifth day of treatment in ICU. Blood and urine samples were collected from the controls, as well.

### Outcomes

Outcomes of interest were; transmission of the SARS-CoV-2 into the urine, incidence of acute kidney injury (AKI) in COVID-19 patients, effects of SARS-CoV-2 on kidney injury markers including serum cystatin C, and urine Neutrophil Gelatinase-Associated Lipocalin (NGAL) and kidney injury molecule-1 (KIM-1) levels. The effects of SARS-CoV-2 on kidney function tests including serum and urine creatinine levels, CKD-EPI (Chronic Kidney Disease Epidemiology Collaboration) creatinine, CKD-EPI cystatin C and CKD-EPI creatinine-cystatin C equations levels, and proteinuria and hematuria were also evaluated. The survival analysis and determining of the potential predictor parameters for specific mortality of the COVID-19 were secondary outcomes of interest.

### Data source/measurement, parameters

The demographic characteristics, laboratory data and medications were extracted from our prospectively noted and/or past medical records. AKI was defined as an increase in serum creatinine by 0.3 mg/dl within 48 hours or a 50% increase in serum creatinine from baseline within 7 days according to the KDIGO criteria [8]. Venous blood samples were collected from the participants to test serum creatinine and cystatin C levels. After the centrifuge of the blood samples for 10 minutes at 4000 g, serum was obtained and serum levels of creatinine and cystatin C were determined using a photometric test on a AU5800 clinical chemistry analyzer (Beckman Coulter Inc. Brea, CA, USA) and nephelometric test on Immage 800 Rate Nephelometer (Beckman Coulter Inc. Brea, CA, USA), respectively. Then, we estimated GFR (eGFR) using the CKD-EPI equations, where Scr is serum creatinine and Scys is serum cystatin C;

*CKD-EPI:* 141 × min(Scr/κ,1)^α^ × max(Scr/κ,1)^−1.209^ × 0.993^Age^ [×1.018 if female] [×1.159 if black], where κ is 0.7 for females and 0.9 for males, α is −0.329 for females and −0.411 for males, min is the minimum of Scr/κ or 1, and max is the maximum of Scr/κ or 1.

*CKD-EPI cystatin C:* 133 × min(Scys/0.8)^−0.499^ × max(Scys/0.8)^−1.328^ × 0.996^Age^ [×0.932 if female], where min indicates the minimum of Scr/κ or 1 and max indicates the maximum of Scys/κ or 1.

*CKD-EPI creatinine-cystatin C:* 135 × min(Scr/κ,1)^α^ × max(Scr/κ,1)^−0.601^ × min(Scys/0.8)^−0.375^ × max(Scys/0.8)^−0.711^ × 0.995^Age^ [×0.969 if female] [×1.08 if black], where κ is 0.7 for females and 0.9 for males, α is −0.248 for females and −0.207 for males, min indicates the minimum of Scr/κ or 1, and max indicates the maximum of Scr/κ or 1.

Single voided morning urine samples and single morning urine samples from urethral catheter were obtained from the patients in stable condition and controls, and patients in ICU, respectively. Spot urine KIM-1 and NGAL levels were determined by enzyme-linked immunosorbent assay (ELISA) using specific Human KIM-1 ELISA kit (SinoGeneClon Biotech, Hangzhou, China) and specific Human NGAL ELISA kit (Thermo Fisher Scientific, Rockford, IL, USA), respectively. Urine creatinine levels were determined using a photometric test on a AU5800 clinical chemistry analyzer (Beckman Coulter Inc. Brea, CA, USA). Spot urine protein and red blood cells (RBC) estimations were performed by FUS-200/H-800 automated urinalysis system with flow cell digital imaging technology (Dirui Industrial Co., Ltd., Changchun, China).

After obtaining of the RT-PCR tests results, the diagnosis of COVID-19 was confirmed for all of the highly suspected cases. The participants divided the two main groups as COVID-19 patients and controls, and then, divided to four final groups as ‘‘COVID-19 patients before treatment’’, ‘‘COVID-19 patients after treatment’’, COVID-19 patients under treatment in ICU’’ and ‘‘controls’’. The parameters age, body mass index (BMI), comorbidity, serum creatinine and cystatin C levels, CKD-EPI, CKD-EPI cystatin C and CKD-EPI creatinine-cystatin C eGFR levels, urine creatinine, KIM-1 and NGAL levels, urine KIM-1/creatinine and NGAL/creatinine ratios were statistically compared between the groups. Urine Ph levels and incidences of proteinuria and microhematuria were also compared.

Based on our comparative statistical results, several covariates were selected for Kaplan-Meier survival and Cox regression analyses. The compared parameters exhibiting p values ≤ 0.1 were considered as covariates. These covariates were age, comorbidities, serum creatinine and cystatin C levels, urine KIM-1/creatinine and NGAL/creatinine levels, and CKD-EPI, CKD-EPI cystatin C and CKD-EPI creatinine-cystatin C eGFR levels.

### Data analysis

Due to the lack of associated data in the literature, a minimum number of participants for sample size could not be calculated and instead of this, a pilot study has been designed. All of the above mentioned parameters of the participants were investigated without any missing data. Statistical analysis was performed with SPSS Version 22.0 statistic software package (IBM SPSS Inc., Chicago, IL). Data distributions and test of normality were evaluated with Shapiro-Wilk test. Descriptive statistic methods (mean±standard deviation and median±interquartile range) were used to evaluate data. We compared the normally distributed and not normally distributed parametric data between the main groups using independent t and Mann-Whitney U tests, respectively. In the analysis of the final groups, we used the One-Way Anova and Kruskal-Wallis tests, respectively. For post-hoc analysis of the One-Way Anova and Kruskal-Wallis tests, we used the Tukey’s and Mann-Whitney U tests, respectively. Chi-square test was also used in the comparison of the nonparametric categorical variables. The associations between covariates including kidney disease indicators and death from COVID-19 were examined using Cox proportional hazard regression analysis.

In multivariable model, we assessed interactions of kidney disease indicators with COVID-19 specific mortality after adjusting the parameters had ≤ 0.1 p values in the univariable analysis results. The adjusted parameters were age, sex and comorbidities. Differences were considered as significant at two-sided P < 0.05 and 95% confidence interval.

## Results

### Demographics and baseline characteristics

A total of 47 participants who met the inclusion criteria were included in the study. Among them, 36 (78.3%) were diagnosed with COVID-19 and remaining were the controls. Of the participants, 26 (55.3%) were male and remaining (44.7%) were female. The mean age of the study cohort was 55.77±17.47 years. Twenty-six participants (55.3%) had comorbid diseases and 15 of them had multiple comorbidity. The mean ages were 59.78±17.11 and 42.64±11.51 years in patients diagnosed with COVID-19 and controls, respectively (p=0.003). In patients with COVID-19, male/female ratio was 18/18, whereas it was 8/3 in the controls (p=0.30). Among the COVID-19 patients, 21 (58.3%) had at least a comorbidity and 13 (36.1%) had multiple comorbidity; whereas 5 (45.5%) of the controls had at least one comorbidity and 2 (18.2%) of them had multiple comorbidity (p=0.50 and p=0.46, respectively) (Table 1). The median duration from onset of symptoms to hospitalization was 3.5 (2-6) days in the COVID-19 patients. The median duration from onset symptoms to ICU admission and from hospitalization to ICU admission were 4 (3-8) and 3 (1-4) days, respectively. The most common symptoms at onset of the disease were fever (n=30, 81.1%), fatigue and myalgia (n=21, 56.7%), and cough (n=17, 46%). The others were diarrhea (n=11, 29.8%), headache and dizziness (n=11, 29.8%), and shortness of breath (n=10, 27%). All of the COVID-19 patients had specific CT findings in their chest CT imaging. All of the patients, except one, in the COVID-19 patients before treatment group were treated in the specific COVID-19 clinic and discharged uneventfully after a median 4.5 (4-9) days of hospital stay. One patient died (7.69%) from COVID-19 on his 5^th^ day of treatment. The median duration of the hospital stay was 5 (5-20) and 13.5 (7-20) days for the COVID-19 patients after treatment group and COVID-19 patients treated in ICU group, respectively. Eleven patients (91.7%) died from COVID-19 in the COVID-19 patients treated in ICU group and one patient was discharged after 15 days treatment in ICU. The entubation rate was 58.3% in the COVID-19 patients treated in ICU. The median time from admission to ICU to entubation and duration of entubation period were 5 (1-9) days and 12 (2-16) days.

**Table 1.** **Demographics, kidney function parameters, kidney damage markers, and urine analyses results of COVID-19 patients and controls**.

|  | **COVID-19 patients**<br>**(n=36)** | **Controls**<br>**(n=11)** | **P** |
| --- | --- | --- | --- |
| **Age (years) (Mean±SD)** | 59.78±17.11 | 42.64±11.51 | 0.03* |
| **BMI (Mean±SD)** | 27.20±2.68 | 27.83±3.10 | 0.51* |
| **Sex (n, %)** |  |  | 0.30** |
| Male | 18, 50 | 8, 72.7 |  |
| Female | 18, 50 | 3, 27.3 |  |
| **Comorbidity (n, %)** |  |  | 0.50** |
| Yes | 21, 58.3 | 5, 45.5 |  |
| No | 15, 41.7 | 11, 54.5 |  |
| (Some participants had multiple) |  |  |  |
| None | 11, 30.6 | 9, 81.2 |  |
| DM | 12, 33.4 | 4, 36.4 |  |
| Hypertension | 15, 41.7 | 2, 18.2 |  |
| CAD | 10, 27.8 | 0, 0.0 |  |
| Others | 2, 5.5 | 0, 0.0 |  |
| **Multiple comorbidity (n, %)** |  |  | 0.46** |
| Yes | 13, 36.1 | 2, 18.2 |  |
| No | 23, 63.9 | 9, 81.8 |  |
| **SARS-CoV-2 positive urine samples (n)** | 2 | 0 |  |
| **Serum Cre (mg/dL) (Median±IQR)** | 0.75±0.39 | 0.76±0.22 | 0.33# |
| **Serum Cyst C (mg/L) (Median±IQR)** | 0.96±0.59 | 0.70±0.17 | 0.003# |
| **CKD-EPI (Mean±SD)** | 86.66±28.02 | 114.72±9.40 | 0.002* |
| **CKD-EPI Cyst C (Mean±SD)** | 86.66±49.01 | 136.33±25.35 | 0.002* |
| **CKD-EPI Cre-Cyst C (Mean±SD)** | 99.57±46.63 | 136.39±24.99 | 0.01* |
| **Urine KIM-1 (ng/mL) (Mean±SD)** | 12.95±5.82 | 10.42±4.02 | 0.18* |
| **Urine KIM-1/Urine Cre (ng/mg) (Median±IQR)** | 15.97±54.24 | 9.31±15.48 | 0.46# |
| **Urine NGAL (ng/mL) (Median±IQR)** | 61.26±75.35 | 21.73±154.60 | 0.11# |
| **Urine NGAL/Urine Cre (ng/mg) (Median±IQR)** | 39.22±126.66 | 12.48±169.50 | 0.09# |
| **Urine Ph (Median±IQR)** | 6.0±0.9 | 6.0±0.0 | 0.17# |
| **Microhematuria (n, %)** |  |  | 0.03** |
| Yes | 16, 44.4 | 1, 9.1 |  |
| No | 20, 55.6 | 10, 81.9 |  |
| **Proteinuria (n, %)** |  |  | 0.004** |
| Yes | 17, 47.2 | 0, 0.0 |  |
| No | 20, 52.8 | 11, 100.0 |  |

COVID-19: Coronavirus disease 2019, BMI: Body mass index, DM: Diabetes mellitus, CAD: Coronary artery disease, Cre: Creatinine, Cyst C: Cystatin C, CKD-EPI: Chronic Kidney Disease Epidemiology Collaboration, KIM-1: Kidney injury molecule 1, NGAL: Neutrophil gelatinase-associated Lipocalin, SD: Standard deviation, IQR: Interquantile range, \* Independent t test, \*\* Chi-square test, # Mann-Whitney U test.

### Urine RT-PCR test, acute kidney injury, and kidney function and damage parameters

SARS-CoV-2 was detected in urine sample just in one COVID-19 patient (2.78%) in our cohort. The patient age was 80’s who had previous hypertension history. His baseline creatine value was 1.61 mg/dL. He exhibited AKI with 2.45 mg/dL serum creatinine level. The highest level of the serum creatinine was detected as 3.59 mg/dL. His kidney injury and function parameters did not show significant differences when compared to other COVID-19 patients under treatment in ICU. The incidence of AKI could not be evaluated in the COVID-19 patients before treatment group, because of the lack of the baseline serum creatinine values. Overall AKI incidence was 16% in the COVID-19 patients. Table 2 provides separate AKI incidences in the COVID-19 patients before treatment and COVID-19 patients in treated in ICU groups. Serum cystatin C levels were detected significantly higher in patients with COVID-19. Moreover, kidney function parameters CKD-EPI, CKD-EPI cystatin C and CKD-EPI creatinine-cystatin C eGFR levels were significantly higher in the COVID-19 patients compared to the controls (Table 1). In the urine analysis, micro-hematuria was detected in the 16 samples and one sample of the patients with COVID-19 and controls, respectively (p=0.03). Similarly, 17 of the COVID-19 patients had proteinuria whereas the controls had any (p=0.004) (Table 1).

**Table 2.** **AKI Incidences in the COVID-19 Patients in our study cohort**.

|  | Pre-diagnosis serum<br>Cre levels (mg/dL) | Serum Cre levels at<br>diagnosis (mg/dL) | Final Serum Cre<br>levels (mg/dL) | Comorbidity |
| --- | --- | --- | --- | --- |
| **COVID-19 patients before treatment group**<br>**AKI Incidence: 2/13, 15.38%** |  |  |  |  |
| Patient 1 | 0.92 | 1.46 | 0.98 | HT |
| Patient 2 | 0.89 | 2.54 | 1.38 | DM |
| **COVID-19 patients treated in ICU group**<br>**AKI Incidence: 2/12, 16.66%** |  |  |  |  |
| Patient 1 | 0.96 | 1.61 | 2.04 | HT |
| Patient 2 | 1.68 | 2.11 | 3.22 | CRF |

Cre: Creatinine, HT: Hypertension, DM: Diabetes mellitus, CRF: Chronic renal failure.

In the comparative analysis of the final groups, the mean ages were 56.46±15.95, 51.00±14.91, 71.42±14.62, and 42.64±11.51 years in the COVID-19 patients before treatment, COVID-19 patients after treatment, COVID-19 patients under treatment in ICU and control groups, respectively (p<0.001). The post hoc analysis revealed that higher mean age of the COVID-19 patients under treatment in ICU accounted for the difference. Serum creatinine and cystatin C levels, urine KIM-1/creatinine levels, and CKD-EPI, CKD-EPI cystatin C and CKD-EPI creatinine-cystatin C eGFR levels exhibited significant difference in the groups (Table 3). The post hoc analysis revealed the causes of the difference were COVID-19 patients before treatment and COVID-19 patients under treatment in ICU groups More altered kidney function and increased acute kidney damage in these patients were responsible for the difference. Additionally, incidences of comorbidity and proteinuria in the urine analysis were higher in the COVID-19 patients under treatment in ICU group (Table 4).

**Table 3.** **Demographics, kidney function parameters, kidney damage markers, and urine analyses results of the participants in the groups**.

|  | COVID-19 patients before treatment (n=13) | COVID-19 patients after treatment (n=11) | COVID-19 patients under treatment in ICU (n=12) | Controls (n=11) | P |
| --- | --- | --- | --- | --- | --- |
| **Age (years) (Mean±SD)** | 56.46±15.95 | 51.00±14.91 | 71.42±14.62 | 42.64±11.51 | <0.001* |
| **BMI (Mean±SD)** | 28.35±2.25 | 27.19±3.50 | 25.96±1.71 | 27.83±3.10 | 0.16* |
| **SARS-CoV-2 positive urine samples (n)** | 2 | 0 | 0 | 0 |  |
| **Serum Cre (mg/dL) (Median±IQR)** | 0.93±0.34 | 0.67±0.23 | 0.83±0.38 | 0.76±0.22 | <0.001# |
| **Serum Cyst C (mg/L) (Median±IQR)** | 1.04±0.69 | 0.80±0.33 | 1.23±0.84 | 0.70±0.17 | <0.001# |
| **CKD-EPI (Mean±SD)** | 74.53±27.59 | 111.45±10.83 | 77.08±25.94 | 114.72±9.40 | <0.001* |
| **CKD-EPI Cyst C (Mean±SD)** | 79.21±47.81 | 121.85±44.00 | 62.47±37.76 | 136.33±25.35 | <0.001* |
| **CKD-EPI Cre-Cyst C (Mean±SD)** | 85.57±39.75 | 141.40±38.69 | 75.27±37.70 | 136.39±24.99 | <0.001* |
| **Urine KIM-1 (ng/mL) (Mean±SD)** | 11.09±6.56 | 12.90±0.11 | 15.00±4.27 | 10.42±4.02 | 0.08* |
| **Urine KIM-1/Urine Cre (ng/mg) (Median±IQR)** | 4.23±5.93 | 6.09±29.32 | 69.58±27.10 | 9.31±15.48 | <0.001# |
| **Urine NGAL (ng/mL) (Median±IQR)** | 47.58±58.64 | 66.21±83.54 | 81.05±93.80 | 21.73±154.60 | 0.44# |
| **Urine NGAL/Urine Cre (ng/mg) (Median±IQR)** | 27.89±54.34 | 25.95±108.63 | 244.28±350.27 | 12.48±169.50 | 0.007# |
| **Urine Ph (Median±IQR)** | 6.0±1.0 | 6.0±0.5 | 5.5±0.5 | 6.0±0.0 | 0.17# |

COVID-19: Coronavirus disease 2019, ICU: Intensive care unit, BMI: Body mass index, Cre: Creatinine, Cyst C: Cystatin C, CKD-EPI: Chronic Kidney Disease Epidemiology Collaboration, KIM-1: Kidney injury molecule 1, NGAL: Neutrophil gelatinase-associated lipocalin, SD: Standard deviation, IQR: Interquartile range, \* One-way Anova test, # Kruskal Wallis test.

**Table 4.** **Categorical demographics and urine parameters of the participants in the groups**.

|  | **COVID-19 patients before treatment (n=13)** | **COVID-19 patients after treatment (n=11)** | **COVID-19 patients under treatment in ICU (n=12)** | **Controls (n=11)** | **p** |
| --- | --- | --- | --- | --- | --- |
| **Sex (n, %)** |  |  |  |  | 0.26* |
| Male | 6, 46.2 | 4, 36.4 | 8, 66.7 | 8, 72.7 |  |
| Female | 7, 53.8 | 7, 63.6 | 4, 33.3 | 3, 27.3 |  |
| **Comorbidity (n, %)** |  |  |  |  | 0.04* |
| Yes | 8, 61.5 | 3, 27.3 | 10, 83.3 | 5, 45.5 |  |
| No | 5, 38.5 | 8, 72.7 | 2, 16.7 | 6, 54.5 |  |
| **Multiple comorbidity (n, %)** |  |  |  |  | 0.12* |
| Yes | 4, 30.8 | 2, 18.2 | 7, 58.3 | 2, 18.2 |  |
| No | 9, 69.2 | 9, 81.8 | 5, 41.7 | 9, 81.8 |  |
| **Microhematuria (n, %)** |  |  |  |  | 0.10* |
| Yes | 5, 38.5 | 4, 36.4 | 7, 58.3 | 1, 9.1 |  |
| No | 8, 61.5 | 7, 63.6 | 5, 41.7 | 10, 90.9 |  |
| **Proteinuria (n, %)** |  |  |  |  | 0.003* |
| Yes | 3, 23.1 | 6, 54.5 | 8, 66.7 | 0, 0.0 |  |
| No | 10, 76.9 | 5, 45.5 | 4, 33.3 | 11, 100.0 |  |

COVID-19: Coronavirus disease 2019, ICU: Intensive care unit, \* Chi-square test.

In the COVID-19 patients before treatment group, median serum creatinine level significantly decreased and mean CKDEPI eGFR level significantly increased at the end of the treatment. On the other hand, nevertheless, these parameters got worse in COVID-19 patients under treatment in ICU (Table 5).

**Table 5.** **Baseline and final levels of the serum creatinine and CKD-EPI in the COVID-19 patients before treatment and under treatment in ICU groups**.

|  | Parameters | Baseline value | Current value** | p |
| --- | --- | --- | --- | --- |
| **COVID-19 patients before treatment** | **Serum Cre (mg/dL)**<br>(Median±IQR) | 0.93±0.34 | 0.89±0.25 | 0.02* |
|  | **CKD-EPI (Mean±SD)** | 74.53±27.59 | 87.00±43.00 | 0.01# |
| **COVID-19 patients under treatment in ICU** | **Serum Cre (mg/dL)**<br>(Median±IQR) | 0.83±0.38 | 1.11±0.55 | 0.003* |
|  | **CKD-EPI (Mean±SD)** | 77.08±28.50 | 60.75±23.54 | 0.003# |

COVID-19: Coronavirus disease 2019, ICU: Intensive care unit, Cre: creatinine, CKD-EPI: Chronic Kidney Disease Epidemiology Collaboration, SD: Standard deviation, IQR: Interquantile range, \*, Wilcoxon test, # Paired t test, \*\* Current value means the after treatment levels in the COVID-19 patients before treatment group, whereas it means the last determined levels during the study period in the COVID-19 patients under treatment in ICU group.

The mean age, median serum cystatin C level, mean CKD-EPI level, and median urine KIM-1/creatinine and urine NGAL/creatinine levels were significantly higher among patients who were died because of COVID-19 compared to survivors of COVID-19 (Table 6). Incidences of comorbid diseases and proteinuria were also significantly higher in died COVID-19 patients.

**Table 6.** **Demographics, kidney function parameters, kidney damage markers, and urine analyses results of survived and died COVID-19 patients**.

|  | **Survived COVID-19 patients (n=24)** | **Died COVID-19 patients (n=12)** | **p** |
| --- | --- | --- | --- |
| **Age (years) (Mean±SD)** | 53.50±15.43 | 72.33±13.23 | 0.001* |
| **BMI (Mean±SD)** | 27.77±2.90 | 26.06±1.74 | 0.17* |
| **Serum Cre (mg/dL) (Median±IQR)** | 0.73±0.32 | 1.01±0.88 | 0.11# |
| **Serum Cyst C (mg/L) (Median±IQR)** | 0.86±0.37 | 1.52±0.66 | 0.01# |
| **CKD-EPI (Mean±SD)** | 93.50±28.90 | 73.00±21.12 | 0.03* |
| **CKD-EPI Cyst C (Mean±SD)** | 101.35±50.46 | 57.28±30.14 | 0.09* |
| **CKD-EPI Cre-Cyst C (Mean±SD)** | 113.21±47.95 | 69.28±31.29 | 0.06* |
| **Urine KIM-1 (ng/mL) (Mean±SD)** | 12.13±6.23 | 14.58±4.73 | 0.24* |
| **Urine KIM-1/Urine Cre (ng/mg) (Median±IQR)** | 5.59±23.93 | 67.94±32.25 | <0.001 |
| **Urine NGAL (ng/mL) (Median±IQR)** | 53.09±68.50 | 81.06±89.88 | 0.14# |
| **Urine NGAL/Urine Cre (ng/mg) (Median±IQR)** | 26.42±73.45 | 244.29±350.27 | <0.001 |

COVID-19: Coronavirus disease 2019, BMI: Body mass index, Cre: Creatinine, Cyst C: Cystatin C, CKD-EPI: Chronic Kidney Disease Epidemiology Collaboration, KIM-1: Kidney injury molecule 1, NGAL: Neutrophil Gelatinase-Associated Lipocalin, SD: Standard deviation, IQR: Interquantile range, \* Independent t test, # Mann-Whitney U test.

### Association of kidney function and damage parameters with COVID-19 specific death

Kaplan-Meier analysis revealed a significantly higher COVID-19 specific death rates for elderly patients and for patients with altered kidney function and abnormal kidney damage markers, including elevated serum cystatin C level, elevated CKD-EPI, CKD-EPI cystatin C and CKD-EPI creatinine-cystatin C eGFR levels, and elevated urine KIM-1/creatinine and urine NGAL/creatinine levels (Figure 1). Univariate Cox regression analysis showed that age above 65 years was associated with COVID-19 specific death. In addition, the kidney function and damage markers including elevated serum cystatin C level, elevated urine KIM-1/creatinine and NGAL/creatinine levels, CKD-EPI eGFR levels, and proteinuria were also associated with COVID-19 specific death (Table 7). After adjusting for age, sex, and comorbidities, the parameters urine KIM-1/creatinine ratio and proteinuria of any degree were associated with COVID-19 specific death (Table 8).

**Table 7.** **Univariable Cox regression analysis of association between abnormal kidney function and kidney damage with the COVID-19 specific death in patients with COVID-19**.

|  |  |  |  |
| --- | --- | --- | --- |
| **Age > 65 years** | 3.08 | 1.001-9.501 | 0.04 |
| **Any comorbidity** | 2.93 | 0.85-10.72 | 0.07 |
| **Cyst C Elevated** | 5.95 | 1.62-21.79 | 0.002 |
| **Urine KIM-1/Cre Elevated** | 6.57 | 1.43-30.02 | 0.004 |
| **Urine NGAL/Cre Elevated** | 29.04 | 0.09-90.06 | 0.06 |
| **CKDEPI Elevated** | 3.81 | 1.04-13.90 | 0.02 |
| **CKDEPI Cyst C Elevated** | 3.99 | 1.09-14.63 | 0.02 |
| **CKDEPI Cre-Cyst C Elevated** | 3.38 | 1.04-11.06 | 0.03 |
| **Proteinuria Any degree** | 3.38 | 1.03-11.04 | 0.03 |

Cyst C: Cystatin C, Cre: Creatinine, KIM-1: Kidney injury molecule 1, NGAL: Neutrophil gelatinase-associated lipocalin, CKD-EPI: Chronic Kidney Disease Epidemiology Collaboration.

**Table 8.** **Age, sex and comorbidities adjusted multivariable Cox regression analysis of association between abnormal kidney function and kidney damage with the COVID-19 specific death in patients with COVID-19**.

|  | **HRs** | **95% CI** | **p value** |
| --- | --- | --- | --- |
| **Cyst C Elevated** | 1.42 | 0.00-2.52 | 0.96 |
| **Urine KIM-1/Cre Elevated** | 6.11 | 1.22-30.53 | 0.02 |
| **Urine NGAL/Cre Elevated** | 3.35 | 0.00-3.44 | 0.96 |
| **CKDEPI Elevated** | 4.44 | 0.88-22.18 | 0.06 |
| **CKDEPI Cyst C Elevated** | 2.10 | 0.33-13.39 | 0.43 |
| **CKDEPI Cre-Cyst C Elevated** | 2.00 | 0.29-13.56 | 0.47 |
| **Proteinuria** | 5.35 | 1.45-19.71 | 0.01 |

Cyst C: Cystatin C, Cre: Creatinine, KIM-1: Kidney injury molecule 1, NGAL: Neutrophil gelatinase-associated lipocalin, CKD-EPI: Chronic Kidney Disease Epidemiology Collaboration.

**Figure 1.**
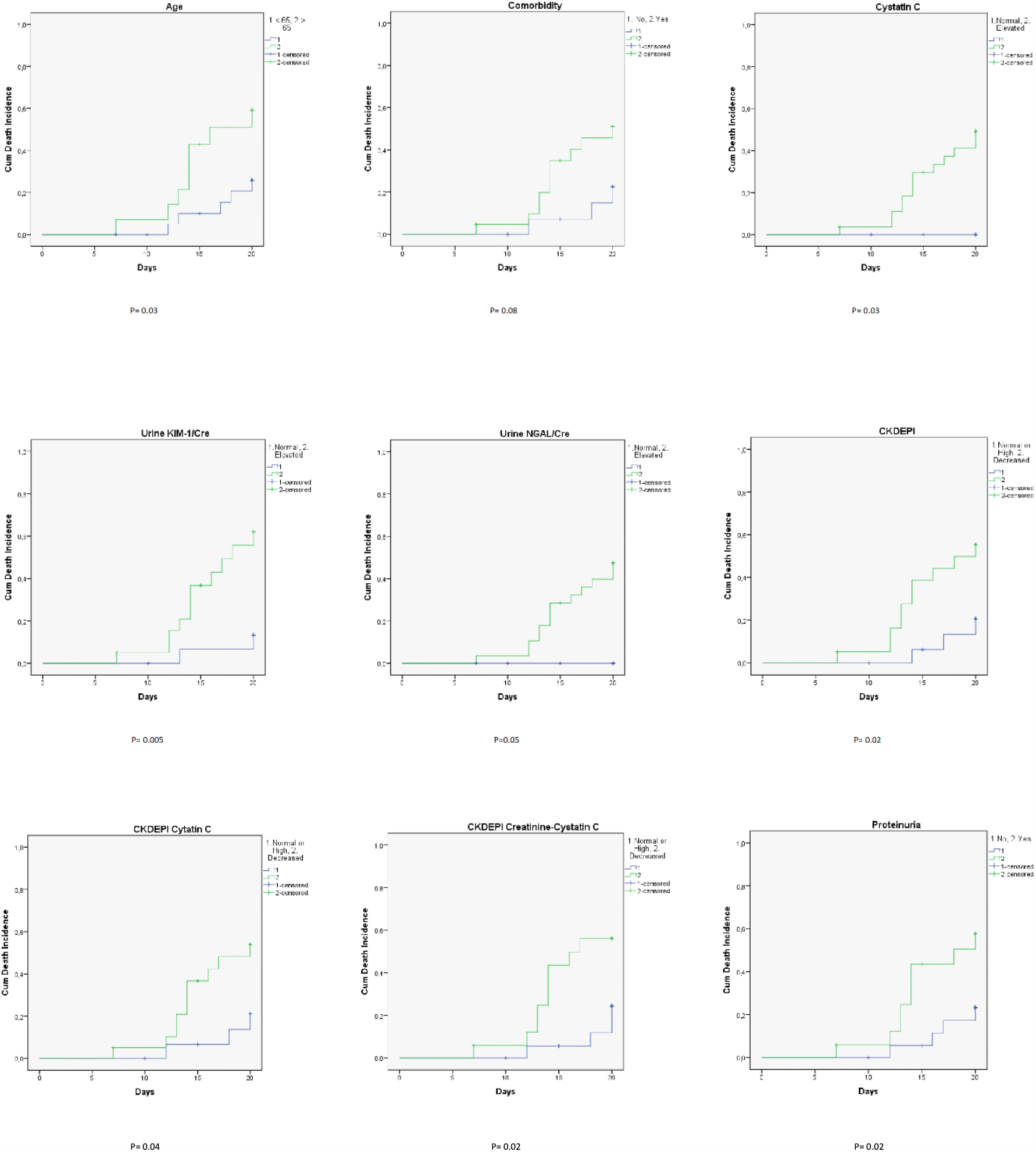
Kaplan-Meier analysis graphics for association of some kidney parameters between COVID-19 specific death rates.

## Discussion

In the recent literature, it is reported that kidney involvement is frequent in COVID-19. Abdominal computerized tomography scan of the COVID-19 patients showed reduced density in the kidneys, indicating the inflammation and oedema [9]. About 40% of the COVID-19 patients may have proteinuria and haematuria on hospital admission [10]. Increased blood urea nitrogen and serum creatinine levels were reported in 14% and 10% of the hospitalized COVID-19 patients. In some of these patients, even if they not diagnosed with AKI at admission, during the hospitalization they could be gradually worsened and diagnosed as AKI [10]. According to the recent literature, AKI incidence in hospitalized COVID-19 patients ranges from 8% to 22% [11].

It is well known that, apart from the alveolar cells in the lungs, many other tissue express ACE2. For instance, the heart, the gut and the kidney has considerable ACE2 expression levels [12]. The National Center for Biotechnology Information’s (NCBI) Gene database reports that, in the human body, the kidney is the fourth highly ACE2 expressing organ following the small intestine, duodenum and gall bladder, respectively [13]. Testis, heart and colon are the other highly ACE2 expressing organs. ACE2 expression level is much higher, nearly 100 fold, in the kidney than that in the respiratory organs [13]. In the kidney, the brush border of proximal tubular cells is the main source of the ACE2 expression. To a lesser extent, podocytes express ACE2. But it is not or scarcely any expressed in glomerular endothelial and mesangial cells [14]. In a recent study based on single-cell transcriptome analysis, Pan et al. [15] investigated 42,589 cells from 15 normal kidney samples and showed that proximal straight tubule cells and podocytes expresses ACE2 highly. However, it is still under debate whether ACE2 expressing kidney is targeted and affected by SARS-CoV-2 infection. On the other side, several current reports showing the co-occurrence of AKI with COVID-19 [9, 11, 16, 17] suggests that SARS-CoV-2 may have a tropism for the kidney. In this regard, a recent study by Diao et al.[18] is quite remarkable. The study showed that one of the specific targets for SARS-CoV-2 is the kidney tissue. The authors found specific SARS-CoV-2 nucleocapsid protein in the kidney specimens and they found that viral antigens accumulated in kidney tubules as a result of postmortem tissue analysis. They concluded that SARS-CoV-2 directly infect human kidney tubules to induce acute tubular damage. In their opinion, beside direct cytotoxicity, it also initiate macrophage and complement mediated tubular pathogenesis secondary to accumulated viral antigens. Similarly, based on their findings, Pan et al. [15] also concluded that the cytopathic effects of SARS-CoV-2 on proximal straight tubule cells and podocytes may cause AKI in patients with COVID-19.

Recent evidences provide that AKI is associated with increased morbidity and mortality in COVID-19 patients. It is considered a marker of disease severity and a negative prognostic factor for survival, as well [11, 17, 19, 20]. Although the possible mechanism of kidney injury by SARS-Cov-2 and COVID-19 associated AKI are well described in the literature, concrete evidence for acute kidney damage in COVID-19 patients is still lacking. Aditionally, available data on COVID-19 associated AKI are reporting incidence on the basis of case series, retrospective studies and a few prospective studies. All of those studies, determined the AKI based on the serum creatinine levels without using accepted acute kidney damage markers [10].

In this prospective pilot study, we investigated the incidence of AKI prospectively in the COVID-19 patients. Moreover, we examined the acute kidney damage markers to obtain a concrete evidence in terms of COVID-19 associated acute kidney damage. Further, we used these markers in addition with the other kidney function parameters including serum creatinine level, eGFR levels and urine analysis in the prediction of COVID-19 specific mortality by using survival analysis. We observed that the incidence of AKI as 16% in hospitalized patients with COVID-19. The incidences were 15.38% and 16.66% in the COVID-19 patients after treatment group and COVID-19 patients under treatment in ICU group, respectively. Due to the lack of the baseline values prior the disease onset, AKI incidence could not determined in the COVID-19 patients before treatment group. We revealed impaired eGFR levels in patients with COVID-19 compared to controls, as well. Additionally, the incidences of micro-hematuria and proteinuria were significantly higher in the COVID-19 patients. Impairment in the kidney function was significantly higher in the as-yet-untreated COVID-19 patients and COVID-19 patients under treatment in ICU. The incidences of micro-hematuria and proteinuria were also higher in those patients. These findings support that COVID-19 affects kidney functions adversely and proportionally with severity of the disease. Besides, we showed the considerable recovery in kidney function with the treatment of COVID-19.

This is the first study investigating the kidney damage markers in COVID-19 patients. We found that COVID-19 patients under treatment in ICU exhibited extremely higher levels of serum cystatin C, and urine KIM-1/creatinine and urine NGAL/creatinine ratios. These results clearly described the acute kidney damage by COVID-19 using molecular kidney damage markers for the first time in the literature. Lowered CKD-EPI, CKD-EPI cystatin C and CKD-EPI creatinine-cystatin C eGFR levels were determined in them, as well.

Currently, it has been reported that kidney disease on admission and AKI during hospitalization were associated with an increased risk of in-hospital death [17]. In our study, indicators of altered kidney function and kidney damage were significantly higher in died COVID-19 patients compared the survived ones. Our univariable Cox regression model indicated that kidney function parameters and damage markers could predict the survival outcomes of COVID-19 patients. After adjusting for age, gender and comorbidity, we found that urine KIM-1/creatinine ratio and proteinuria were associated with COVID-19 specific death. In this regard, considering kidney function and kidney damage markers must not be ignored in the COVID-19 patients, and serial monitoring of them should be considered. Moreover, impairment in the kidney function and/or emerging kidney damage should alert clinicians during the management of COVID-19 patients.

The present study has several limitations. First, our cohort has small number of patients. However, we intended to obtain our pilot findings about COVID-19 associated functional impairment in the kidney and kidney damage in default of sufficient data in the literature. Second, even we performed adjusted analysis, other parameters and covariates than our investigated might have played a role on survival analysis. In other respects, our study has some strengths. One of them is, serial monitoring of the serum creatinine levels with available accurate baseline values. Second one is the use of molecular kidney damage markers which could indicate functional alteration in the kidney prior the elevation of serum creatinine levels. This provided us to clearly describe the acute kidney damage by COVID-19 for the first time in the literature. Classification of the COVID-19 patients as before treatment, after treatment and under treatment in ICU might have provided more accurate findings, therefore, it may be accepted as other strength. Finally, this is the first study investigating the predictive role of kidney damage markers in determining the survival outcomes of the patients with COVID-19.

## Data Availability

Research data are not shared.

## Acknowledgements

This work was supported by a grant from Health Institutes of Turkey (TUSEB) with a grant number of 2020-CV-01-8708/8969. We thank administration and staffs of the TUSEB. We would like to express our special thanks of gratitude to our clinic nurses for their substantial contribution to the study while they exhibited heroic efforts during the fight against COVID-19. We want to thank our allied health personnel for their infinite pains, as well.

## Data availability statement

Research data are not shared.

## Notes

### Competing Interest Statement

Mustafa Zafer Temiz, Ibrahim Hacibey, Ramazan Omer Yazar, Mehmet Salih Sevdi, Suat Hayri Kucuk, Levent Doganay, Muhammet Murat Dincer, Kerem Erkalp, and Ahmet Yaser Muslumanoglu were granted by Health Institutes of Turkey (TUSEB) with a grant number of 2020.CV.01.8708/8969.

### Author Declarations

The Institutional Review Board (Bagcilar Training and Research Hospital Local Ethic Committee, Licenced by Turkish Drug and Medical Device Foundation) approved the study (Approval No: 2020.05.1.06.038). The principles outlined in the Declaration of Helsinki were followed and informed consent from participants was obtained, as well. Approval of Turkish Ministry of Health also provided for the study.

